# Exploring the factors associated with prelacteal feeding in Papua New Guinea: a population-based cross-sectional study

**DOI:** 10.1101/2024.06.18.24309085

**Authors:** McKenzie Maviso, Elias Namosha, Georgia Guldan

**Affiliations:** Division of Public Health, School of Medicine and Health Sciences, University of Papua New Guinea, Port Moresby, Papua New Guinea

**Author notes:** Correspondence to McKenzie Maviso.

## Abstract

**Objective:** Prelacteal feeds disrupt early breastfeeding initiation and exclusive breastfeeding and increase the risk of childhood illnesses and under-five mortality. Despite its negative health outcomes, prelacteal feeding prevails in Papua New Guinea (PNG). This study aimed to investigate the factors associated with prelacteal feeding among women in PNG.

**Design and setting:** A population-based cross-sectional study based on Demographic and Health Survey data

**Setting:** Papua New Guinea

**Participants:** A total weighted sample of 4399 women was included in the study.

**Main outcome measures:** Prelacteal feeding, modeled using multivariable logistic regression.

**Results:** About 10% (95% CI: 9 to 11) of women provided prelacteal feeds to their infants in PNG. The most frequently reported prelacteal feed was plain water (71.7%), followed by grains (eg, noodles) (47.1%), dark green leafy vegetables (42.1%), and soup (39.7%). Women with no formal (AOR 1.4, 95% CI: 1.0 to 3.0) or primary education (AOR 1.5, 95% CI: 1.0 to 2.9), from the Islands region (AOR 2.3, 95% CI: 1.5 to 3.5), who had a cesarean section (AOR 4.1, 95% CI: 2.4 to 7.2), and who had given birth at home or in the village (AOR 3.7, 95% CI: 2.1 to 6.8) had higher odds of providing prelacteal feeds. No statistically significant association was found between immediate newborn skin-to-skin contact after birth and prelacteal feeding.

**Conclusion:** Our study underscores the role of sociodemographic and healthcare system factors in prelacteal feeding. Strengthening healthcare providers’ capacity to increase mothers’ awareness of optimal breastfeeding and promote more health facility births is warranted. Comprehensive breastfeeding education should also be promoted at antenatal clinics and during outreach healthcare activities.

**Strengths and limitations of the study:**

- The rigor of the methodological approach and representativeness of the survey render its results and recommendations generalisable to the country.
- The study’s findings, based on appropriate estimation adjustments and sample design, are representative of female population in PNG; thus could guide policy development and design for effective infant and child feeding interventions.
- The study’s cross-sectional nature means that a temporal relationship cannot be established between the outcome and independent variables.
- Prelacteal feeding was assessed based on the maternal self-report, which might have resulted in desirability bias; mothers may be less likely to report that they have provided prelacteal feeds while knowing its adverse outcomes.
- The DHS did not collect some important information, such as maternal beliefs, misconceptions, knowledge, attitudes, and practices towards breastfeeding, which could impact prelacteal feeding in this context.

## Introduction

Breastfeeding and human milk are the normative standards for infant feeding and nutrition, offering numerous short-and long-term health benefits for both mother and child [1]. Improving breastfeeding rates could potentially save over 800 000 children under five globally every year [2]. Breastfeeding has been proven to decrease childhood diseases such as respiratory tract infections, otitis media, diarrheal diseases, undernutrition, and childhood obesity [2–5]. Also, breastfeeding has the potential to prevent approximately 20 000 deaths from various diseases such as breast cancer, diabetes, obesity, and cardiovascular diseases [2,4]. Thus, early initiation of breastfeeding within the first hour after birth, exclusively until six months, and continuing up to two years of age is highly recommended [6].

Despite the multiple benefits of breastfeeding, not all infants worldwide receive early, exclusive, or continued breastfeeding. Only 37% of infants under five are exclusively breastfed in low- and middle-income countries (LMICs) [2,7], while inappropriate feeding practices account for two-thirds of malnutrition-related deaths during the first 24 months of life [8]. Delays in breastfeeding initiation within the first hour of birth can disrupt exclusive breastfeeding and increase the odds of prelacteal feeding, including neonatal mortality [9,10].

Prelacteal feeding, which is the provision of any food or liquid to an infant before breastfeeding is established during the first three days [11], has been shown to interrupt breastfeeding initiation and impede exclusive breastfeeding [12–14]. It can also be detrimental to breastfeeding and harmful to health, as it deprives infants of valuable nutrients and antibody-rich colostrum and exposes them to the risk of infection [15–18]. Studies have shown that the type of prelacteal feed given to the newborn is predominantly associated with sociodemographic and economic factors (eg, education, occupation, income), maternal health care service utilization, mode of birth, place of giving birth, breastfeeding education, and unskilled birth attendants [16,19–21]. Other factors such as poor breastfeeding knowledge, breastfeeding problems, maternal illness, delayed initiation of breastfeeding, and social and cultural norms are factors associated with prelacteal feeding [22–25].

Additionally, prelacteal feeding predisposes infants to pathogenic factors, creating physiological disruptions in the immature gastrointestinal systems, leading to infantile diarrhoea, and affecting infants’ development [18,26]. However, despite its consequences, it remains a prevailing nutritional malpractice and a risk indicator for infant morbidity and mortality in LMICs [27]. A population-based analysis of 57 participating countries found that prelacteal feeding remained prevalent in LMICs, with Asia having the highest prevalence (nearly 60%) compared to other regions (<40%) [28]. Similarly, recent studies in the Asia-Pacific, such as Bangladesh, Nepal, and Pakistan, showed high prelacteal feeding rates (29%, 30.2%, and 32%, respectively) [29–31]. Such forms of supplementation can be particularly deleterious in settings where poverty or resource scarcity can impede nutritional potential.

PNG, a linguistically diverse country, has a strong breastfeeding culture, with an estimated breastfeeding rate ranging from 62% to 85% among children aged less than 24 months [32,33]. Prior studies further indicated a high prevalence of early initiation of breastfeeding in the remote settings of Madang (36%) and Gulf (69%) provinces [34,35]. Despite this evidence, a recent DHS finding revealed that 40% of women in PNG delayed breastfeeding initiation within the first hour after birth [36]. Similarly, only about 50% of children aged 6 to 8 months from urban and rural locations in six provinces were fed complementary foods in the country [33]. Suboptimal breastfeeding practices have been associated with maternal age, lack of breastfeeding knowledge and support, poor antenatal education, place and type of birth, poor skin-to-skin contact, and sociocultural norms [25,34,36–38].

There is therefore evidence that suboptimal breastfeeding and the introduction of prelacteal feeds are common in PNG [25,34,37,38]. However, little is known about factors that may influence prelacteal feeding at the population level. There is also insufficient evidence to determine whether PNG women are aware of the risks associated with prelacteal feeding, such as gastroenteritis and respiratory infections. To bridge the knowledge gap and create successful breastfeeding-targeted interventions, further research is required. Hence, this study aimed to assess factors associated with prelacteal feeding in PNG. The study’s results could offer valuable insights for developing innovative strategies to enhance infant and young child feeding and health in the country.

## Methods

### Data source and sampling

This study used a child dataset from the PNGDHS, a cross-sectional, two-stage cluster sampling study representing the entire country from October 2016 to December 2018. The survey used the census units (CU) list from the 2011 PNG National Population and Housing Census as the sampling frame. Stratification of the sample was achieved by two categories: urban and rural areas, resulting in 43 sampling strata, except for the National Capital District, which had no rural areas. Samples of CUs were selected independently in each stratum in two stages. The first stage involved the selection of 800 CUs, which was done through probability proportional to CU size. The second stage involves the selection of 24 households from each cluster through probability random sampling, resulting in a total sample size of approximately 19200 households. The selected sample consisted of 17505 households. Of the 18175 women identified in the interview households, 15198 completed the interviews (84% response rate). The survey used three sets of validated questionnaires to collect data: a household questionnaire, a woman’s questionnaire, and a man’s questionnaire. The dataset used in this study included all the relevant information on child health from all three questionnaires. Furthermore, the study population for this study was restricted to mothers who had given birth in the last three years to enable comparability with the existing literature and minimize recall bias [39]. Women who had never been pregnant were excluded from the analysis. A total weighted sample of 4399 women with complete information about the variables of interest was included in the final analysis. Details of methodology, sampling design, pretesting, training of field workers, and household selection are in the final report [32].

### Study variables

The outcome variable for this study was prelacteal feeding. Women were asked during the survey: “In the first three days after birth, was (child’s name) given anything to drink or eat other than breast milk?” Their responses were coded as “1” for “yes” if the child was given anything other than breast milk or “0” for “no” if prelacteal feeds were not given. Prelacteal feeding is described as any food or liquid given before the onset of lactogenesis II, that is, the onset of copious breast milk secretion occurring within four days of birth [40]. Infant feeding included plain water as the most commonly reported, juices, powdered milk (non-breast milk), infant formula, fruits (papaya and mango), meat products (eggs, chicken, lamb, and fish), vegetables and tubers (potatoes and cassava), and others [32].

The independent variables were sociodemographic characteristics, which included maternal age, marital status, education status, occupation, wealth index, region, and place of residence, and frequency of mass media access, which included reading a newspaper or magazine, listening to the radio, and watching television. Maternal health service use variables included the number of antenatal visits in pregnancy, mode of birth, and place of birth. Child-related factors comprised the child’s birth order, perceived birth size, birth weight, initiation of breastfeeding, and newborn skin-to-skin contact immediately after birth.

### Statistical analysis

Analyses were limited to participants with complete data for the variables of interest using IBM SPSS for Windows, Version 26.0 (IBM Corp., Armonk, NY, USA). The sample was weighted using the primary sampling unit variable, stratification variable, and weight variable to restore its representativeness and get a better estimate throughout the analysis. Descriptive statistics were used to analyze the sociodemographic characteristics of participants, while inferential statistics were employed for generalization to the entire population through bivariate and multivariate analysis. All the variables, whether significant in the chi-square test or not were included in a multivariable analysis to determine their collective associations with HIV testing. All the analyses were performed using the Complex Sample Analysis procedure which was deemed necessary to adjust for sample weight, and multi-stage sampling [41]. Adjusted odds ratios (AORs) with 95% confidence intervals (CIs) were reported. A p ≤ 0.05 was considered statistically significant.

### Patient and public involvement

Patients and the public were not involved in the study.

## Results

### Characteristics of the participants

**Table 1** presents the characteristics of the participants. A total of 4399 women were included in the study. The mean maternal age of the respondents was 28.9 years (SD = 6.9). About half (50.4%) had completed primary education, and most were not working (69.8%). In terms of mass media exposure, over two-thirds of women never read a newspaper or magazine (67.2%), listened to the radio (69.7%), or watched television (80.2%).

**Table 1.** Characteristics of the participants, PNG, 2016–2018 (N = 4399)

| Characteristics | Frequency (n) | Weighted (%) |
| --- | --- | --- |
| Age group (years) |  |  |
| 15–24 | 1323 | (30.1) |
| 25–34 | 2071 | (47.1) |
| 35–44 | 910 | (20.7) |
| 45–49 | 94 | (2.1) |
| Mean age (SD)<sup>†</sup> 28.9 (6.89) |  |  |
| Marital status |  |  |
| Never married | 159 | (3.6) |
| Married | 3984 | (90.6) |
| Separated/divorced | 257 | (5.8) |
| Educational level |  |  |
| No formal education | 1140 | (25.9) |
| Primary | 2218 | (50.4) |
| Secondary | 911 | (20.7) |
| College/tertiary | 130 | (3.0) |
| Occupation |  |  |
| Not working | 3071 | (69.8) |
| Working | 1328 | (30.2) |
| Wealth index |  |  |
| Poorest | 961 | (21.9) |
| Poorer | 900 | (20.5) |
| Middle | 911 | (20.7) |
| Richer | 881 | (20.0) |
| Richest | 745 | (16.9) |
| Frequency of reading a newspaper/magazine<br>(missing, n = 32) |  |  |
| Not at all | 2934 | (67.2) |
| Less than once a week | 772 | (17.7) |
| Once a week | 661 | (15.1) |
| Frequency of listening to the radio (missing, n = 32) |  |  |
| Not at all | 3043 | (69.7) |
| Less than once a week | 769 | (17.6) |
| Once a week | 555 | (12.7) |
| Frequency of watching television (missing, n = 26) |  |  |
| Not at all | 3505 | (80.2) |
| Less than once a week | 393 | (9.0) |
| Once a week | 476 | (10.9) |
| Place of Residence |  |  |
| Urban | 497 | (11.3) |
| Rural | 3902 | (88.7) |
| Region |  |  |
| Southern | 900 | (20.5) |
| Highlands | 1655 | (37.6) |
| Momase | 1192 | (27.1) |
| Islands | 653 | (14.8) |

<sup>†</sup>SD = Standard deviation

### Maternal and child-related factors

**Table 2** presents maternal and child-related factors. The majority of women had at least one antenatal visit during their pregnancies (72.8%) and gave birth in a public health facility (59.6%). Regarding child-related factors, over one-third of the infants (38.1%) had a birth order of four or more. Most weighed ≥2500 g at birth (82.7%) and also half (46.2%) were introduced immediately to newborn skin-to-skin contact after birth with their mothers. About a quarter (25.1%) delayed initiation of breastfeeding for more than an hour of birth.

**Table 2.** Maternal and child-related factors, PNG, 2016–2018 (N = 4399)

| Characteristics | Frequency (n) | Weighted (%) |
| --- | --- | --- |
| Antenatal visits (at least 1) (missing, n = 55) |  |  |
| No | 1182 | (27.2) |
| Yes | 3163 | (72.8) |
| Mode of birth |  |  |
| Vaginal | 4276 | (97.2) |
| Cesarean section | 124 | (2.8) |
| Place of birth |  |  |
| Home/village | 1779 | (40.4) |
| Public health facility | 2621 | (59.6) |
| Birth order |  |  |
| 1<sup>st</sup> | 1,020 | (23.2) |
| 2<sup>nd</sup> | 901 | (20.5) |
| 3<sup>rd</sup> | 801 | (18.2) |
| 4<sup>th</sup> or more | 1677 | (38.1) |
| Birth size (missing, n = 148) |  |  |
| Large | 1578 | (37.1) |
| Average | 1800 | (42.3) |
| Small | 873 | (20.5) |
| Birth weight (g) (missing, n = 2,145) |  |  |
| ≤ 2,500 | 390 | (17.3) |
| ≥ 2,500 | 1864 | (82.7) |
| Newborn skin-to-skin contact (missing, n = 62) |  |  |
| No | 2332 | (53.8) |
| Yes | 2005 | (46.2) |
| Initiation of breastfeeding (missing, n = 968) |  |  |
| Early (<1 hr.) | 2570 | (74.9) |
| Delayed (>1 hr.) | 861 | (25.1) |

### Prevalence of prelacteal feeding

**Table 3** shows the proportion of the type of prelacteal feeds given to newborn infants in PNG. A total of 433 [9.6% (95% CI: 9 to 11)] weighted proportion] women introduced prelacteal feeds to their infants other than breastfeeding in the first three days after the birth. The most frequently reported prelacteal feed was plain water (71.7%), followed by grains (eg, noodles) (47.1%), dark green leafy vegetables (42.1%) and soup (39.7%).

**Table 3.** Types of prelacteal feeds given to newborn infants, PNGDHS 2016–2018.

| Prelacteal feeds | Frequency (n = 1988) |  |  |  |
| --- | --- | --- | --- | --- |
|  | No | (%) | Yes | (%) |
| Baby formula | 1902 | (95.7) | 86 | (4.3) |
| Beans, peas, lentils, nuts | 1629 | (81.9) | 359 | (18.1) |
| Dairy products (eg, cheese, yogurt, other milk products) | 1870 | (94.1) | 118 | (5.9) |
| Dark green leafy vegetables | 1152 | (57.9) | 836 | (42.1) |
| Eggs | 1583 | (79.6) | 405 | (20.4) |
| Fruits (eg, mangoes, papayas, other vitamin A fruits) | 1361 | (68.5) | 627 | (31.5) |
| Gave child juice | 1436 | (72.2) | 552 | (27.8) |
| Grains (eg, bread, noodles) | 1051 | (52.9) | 937 | (47.1) |
| Liver, heart, other organs | 1881 | (94.6) | 107 | (5.4) |
| Meat (beef, pork, lamb, chicken, fish) | 1699 | (85.5) | 289 | (14.5) |
| Other liquid | 1466 | (73.7) | 522 | (26.3) |
| Other solid-semisolid food | 1405 | (70.7) | 583 | (29.3) |
| Plain water | 563 | (28.3) | 1425 | (71.7) |
| Soup/clear broth | 1199 | (60.3) | 789 | (39.7) |
| Tinned, powdered or fresh milk | 1815 | (91.3) | 173 | (8.7) |
| Tubers (eg, potatoes, cassava) | 1257 | (63.2) | 731 | (36.8) |
| Vegetables (eg, pumpkin, carrots, squash) | 884 | (44.5) | 1104 | (55.5) |

### Factors associated with prelacteal feeding in PNG

**Table 4** shows factors associated with prelacteal feeding in PNG. In the bivariate analysis, educational level, occupation, wealth index, frequency of mass media access (eg, newspaper, radio, and television), region, place of residence, mode of birth, place of birth, and birth weight were all statistically significantly associated with prelacteal feeding. Women with no formal (AOR 1.4; 95% CI: 1.0 to 3.0) or primary education (AOR 1.5, 95% CI: 1.0 to 2.9), from the Islands region (AOR 2.3, 95% CI: 1.5 to 3.5), had a cesarean section (AOR 4.1, 95% CI: 2.4 to 7.2), and who had given birth at home or in the village (AOR 3.8, 95% CI: 2.1 to 6.8) had higher odds of providing prelacteal feeds. However, secondary education (AOR = 0.9, 95% CI: 0.5 to 1.8), the Highlands (AOR 0.9, 95% CI: 0.6 to 1.4) and the Momase region (AOR 0.6, 95% CI: 0.4 to 1.1) were protective factors of prelacteal feeding. No statistically significant association was found between immediate newborn skin-to-skin contact after birth and prelacteal feeding (p = 0.772).

**Table 4.** Multivariate analysis of factors associated with prelacteal feeding in PNG, 2016–2018 (N = 4399)

| Characteristics | Prelacteal feeds |  | COR (95% CI) | AOR (95% CI) | P value* |
| --- | --- | --- | --- | --- | --- |
|  | No, n = 3967 (%) | Yes, n = 433 (%) |  |  |  |
| Age group (years) |  |  |  |  | 0.628 |
| 15–24 | 1215 (30.6) | 108 (24.9) | Ref. | Ref. |  |
| 25–34 | 1858 (46.8) | 213 (49.2) | 1.3 (0.9 to 1.8) | 1.3 (0.8 to 1.9) |  |
| 35–44 | 806 (20.3) | 104 (24.1) | 1.5 (1.0 to 2.0) | 1.3 (0.8 to 2.2) |  |
| 45–49 | 87 (2.2) | 8 (1.8) | 1.0 (0.4 to 2.5) | 0.9 (0.4 to 2.8) |  |
| Marital status |  |  |  |  | 0.595 |
| Never married | 141 (3.6) | 18 (4.2) | 1.4 (0.7 to 2.8) | 1.5 (0.7 to 3.1) |  |
| Married | 3591 (90.5) | 393 (90.8) | 1.2 (0.7 to 1.9) | 1.3 (0.8 to 2.1) |  |
| Separated/divorced | 235 (5.9) | 22 (5.0) | Ref. | Ref. |  |
| Educational level |  |  |  |  | **0.031** |
| No formal education | 1058 (26.7) | 82 (19.0) | 0.6 (0.3 to 1.1) | 1.4 (1.0 to 3.0) |  |
| Primary | 1976 (49.8) | 242 (56.0) | 0.9 (0.5 to 1.7) | 1.5 (1.0 to 2.9) |  |
| Secondary | 819 (20.6) | 92 (21.3) | 0.8 (0.4 to 1.6) | 0.9 (0.5 to 1.8) |  |
| College/tertiary | 114 (2.9) | 16 (3.7) | Ref. | Ref. |  |
| Occupation |  |  |  |  | 0.382 |
| Not working | 2794 (70.4) | 277 (64.1) | 0.8 (0.6 to 1.0) | 0.9 (0.6 to 1.2) |  |
| Working | 1173 (29.6) | 155 (35.9) | Ref. | Ref. |  |
| Wealth index |  |  |  |  | 0.855 |
| Poorest | 888 (22.4) | 73 (16.9) | 0.6 (0.4 to 0.9) | 1.4 (0.6 to 3.1) |  |
| Poorer | 821 (20.7) | 79 (18.2) | 0.8 (0.5 to 1.1) | 1.5 (0.7 to 3.2) |  |
| Middle | 830 (20.9) | 82 (18.9) | 0.8 (0.5 to 1.2) | 1.4 (0.7 to 2.8) |  |
| Richer | 768 (19.4) | 114 (26.3) | 1.2 (0.8 to 1.7) | 1.4 (0.7 to 2.6) |  |
| Richest | 660 (16.6) | 85 (19.6) | Ref. | Ref. |  |
| Frequency of reading a newspaper/magazine<br>(missing, n = 34) |  |  |  |  | 0.358 |
| Not at all | 2789 (70.9) | 255 (59.0) | Ref. | Ref. |  |
| Less than once a week | 670 (17.0) | 99 (22.9) | 1.6 (1.2 to 2.2) | 1.4 (0.9 to 2.1) |  |
| Once a week | 477 (12.1) | 78 (18.1) | 1.8 (1.3 to 2.6) | 1.2 (0.7 to 2.0) |  |
| Frequency of listening to the radio (missing, n = 49) |  |  |  |  | 0.212 |
| Not at all | 2693 (68.5) | 242 (56.0) | Ref. | Ref. |  |
| Less than once a week | 674 (17.1) | 97 (22.5) | 1.6 (1.2 to 2.3) | 1.2 (0.8 to 1.9) |  |
| Once a week | 567 (14.4) | 93 (21.5) | 1.8 (1.3 to 2.7) | 1.5 (0.9 to 2.4) |  |
| Frequency of watching television (missing, n = 47) |  |  |  |  | 0.444 |
| Not at all | 3189 (80.9) | 317 (73.2) | Ref. | Ref. |  |
| Less than once a week | 344 (8.7) | 49 (11.3) | 1.4 (0.9 to 2.2) | 1.2 (0.7 to 1.9) |  |
| Once a week | 409 (10.4) | 67 (15.5) | 1.6 (1.2 to 2.3) | 1.4 (0.8 to 2.4) |  |
| Region |  |  |  |  | **<0.001** |
| Southern | 810 (20.4) | 91 (21.0) | Ref. | Ref. |  |
| Highlands | 1519 (38.3) | 135 (31.1) | 0.8 (0.6 to 1.1) | 0.9 (0.6 to 1.4) |  |
| Momase | 1103 (27.8) | 89 (20.6) | 0.7 (0.4 to 1.2) | 0.6 (0.4 to 1.1) |  |
| Islands | 535 (13.5) | 118 (27.3) | 1.9 (1.3 to 2.9) | 2.3 (1.5 to 3.5) |  |
| Place of Residence |  |  |  |  | 0.081 |
| Urban | 423 (10.7) | 74 (17.1) | Ref. | Ref. |  |
| Rural | 3541 (89.3) | 358 (82.9) | 0.6 (0.3 to 0.9) | 0.5 (0.2 to 1.1) |  |
| Antenatal visits during pregnancy (at least 1 or more) (missing, n = 69) |  |  |  |  | 0.278 |
| No | 1083 (27.6) | 99 (23.5) | 0.8 (0.6 to 1.1) | 0.8 (0.5 to 1.2) |  |
| Yes | 2840 (72.4) | 323 (76.5) | Ref. | Ref. |  |
| Mode of birth |  |  |  |  | <0.001 |
| Vaginal | 3875 (97.7) | 401 (92.6) | Ref. | Ref. |  |
| Cesarean section | 92 (2.3) | 32 (7.4) | 3.4 (1.9 to 5.7) | 4.1 (2.4 to 7.2) |  |
| Place of birth |  |  |  |  | 0.001 |
| Home/village | 1588 (40.0) | 190 (44.0) | 1.2 (0.9 to 1.5) | 3.7 (2.1 to 6.8) |  |
| Health facility | 2378 (60.0) | 242 (56.0) | Ref. | Ref. |  |
| Birth order |  |  |  |  | 0.219 |
| 1<sup>st</sup> | 918 (23.1) | 102 (23.6) | Ref. | Ref. |  |
| 2<sup>nd</sup> | 833 (21.0) | 68 (15.7) | 0.7 (0.5 to 1.1) | 0.63 (0.4 to 1.0) |  |
| 3<sup>rd</sup> | 721 (18.2) | 80 (18.5) | 0.9 (0.7 to 1.4) | 0.83 (0.5 to 1.3) |  |
| 4<sup>th</sup> or more | 1494 (37.7) | 183 (42.2) | 1.1 (0.8 to 1.5) | 0.91 (0.6 to 1.5) |  |
| Birth size (missing, n = 195) |  |  |  |  | 0.600 |
| Large | 1429 (37.3) | 150 (35.7) | 0.9 (0.7 to 1.3) | 0.8 (0.5 to 1.3) |  |
| Average | 1617 (42.2) | 182 (43.2) | 1.0 (0.7 to 1.4) | 0.9 (0.6 to 1.3) |  |
| Small | 784 (20.5) | 89 (21.1) | Ref. | Ref. |  |
| Birth weight (g) (missing, n = 2145) |  |  |  |  | 0.293 |
| ≤ 2500 | 347 (9.3) | 43 (10.5) | Ref. | Ref. |  |
| ≥ 2500 | 1660 (44.5) | 204 (49.6) | 1.0 (0.7, 1.5) | 1.1 (0.7, 1.8) |  |
| Skin-to-skin contact (missing, n = 85) |  |  |  |  | 0.772 |
| No | 2104 (53.8) | 228 (53.4) | 1.0 (0.7, 1.3) | 0.9 (0.7, 1.3) |  |
| Yes | 1805 (46.2) | 199 (46.6) | Ref. | Ref. |  |
| Initiation of breastfeeding (missing, n = 968) |  |  |  |  | 0.466 |
| Early (< 1 hr.) | 2341 (75.3) | 230 (71.0) | Ref. | Ref. |  |
| Delayed (> 1 hr.) | 767 (24.7) | 94 (29.0) | 1.3 (0.9 to 1.8) | 0.9 (0.6 to 1.3) |  |
| *The significance level is set at 0.05.<br>COR = crude odds ratio; AOR = adjusted odds ratio; Ref. = Reference category. |  |  |  |  |  |

## Discussion

In this study, we assessed the prevalence and factors associated with prelacteal feeding in PNG using nationally representative survey data. Overall, one in ten (10%) women had ever introduced prelacteal feeds to their newborn infants. Our finding is similar to the proportion found in studies conducted in East Africa (12%) [21] and Ethiopia (14%) [16,42], but much lower than findings found in studies conducted in other Asian LMICs (33.9%) [17], including Bangladesh (29%) [31], Nepal (30.6%) [29], and Indonesia (45%) [43]. Reasons for the differences in the rates of prelacteal feeding may be due to sociocultural factors, geographical settings, maternal health service use or availability, and access to media and information across different countries.

### Multivariate analysis of factors associated with prelacteal feeding in PNG, 2016–2018 (N = 4399)

We found a statistically significant association between education level and prelacteal feeding. Women with low educational levels were more likely to practice prelacteal feeding compared to those who were more educated. This finding is consistent with previous studies in sub-Saharan Africa [44] and Indonesia [45] that reported low maternal education as a major determinant of some suboptimal breastfeeding practices. Studies have shown that women who provide prelacteal feeds often come from disadvantaged, poor, and less educated backgrounds [45,46]. Targeted educational and literacy support programs for women with no formal education remain the fundamental prerequisite for imparting breastfeeding and infant feeding information, fostering women’s self-confidence, and enhancing the ability to utilize such information effectively [47,48]. This suggests that while formal education is necessary for all women, short-term basic nutrition education and literacy programs will significantly impact women. Therefore, stakeholders and public health institutions are obligated to ensure the provision of affordable, accessible, and family-focused nutrition, and health education programs for these populations.

Our study found significant variations in prelacteal feeding rates across four major administrative regions of PNG, ranging from 20.6% in the Momase region to 31.2% in the Highlands region. This study further found that women from the Islands region were twice as likely to practice prelacteal feeding compared to those from the Southern region. This could have been influenced by geographic variations (ie, many scattered small islands and atolls), remoteness and lack of health service delivery, and limited access to health information for women. Another possible reason could be the inequitable distribution and allocation of health infrastructure and healthcare services. Access to health services and information is more accessible in some parts of the country, such as the Southern region, surrounding the more centrally located capital, Port Moresby, compared to those in the more distant and peripheral Islands region, which could have contributed to optimal infant feeding practices [36]. The differences in the ethnic composition of different regions and their respective sociocultural norms impacting prelacteal feeding may further offer an alternative explanation [49]. Efforts to improve breastfeeding outcomes should be conceived to consider novel and context-specific strategies for women in remote and isolated settings.

Regarding the mode of birth, women who delivered by cesarean section were four times more likely to practice prelacteal feeding compared to those who delivered vaginally. These findings align with previous studies conducted elsewhere [16,20,21,43,49], indicating that cesarean sections have been recognised as a risk factor for prelacteal feeding. There are numerous reasons suggesting that maternal recovery from pain, immobilisation, and fatigue following a cesarean section may make mothers struggle with breastfeeding. Maternal recovery from pain, immobilisation, and fatigue post-caesarean section are some of the reasons that may cause difficulties in breastfeeding [21,43]. Another possible explanation is that the post-anaesthesia effect may hinder early breastfeeding initiation, causing mothers to introduce prelacteal feeds instead of breast milk [20,49]. This study suggests that healthcare professionals and birthing facilities may be lacking in the necessary breastfeeding support strategies for caesarean mothers. More education and support are also required for infant feeding and the potential challenges women may encounter following caesarean births.

Existing studies show significant relationships between home or village birth and prelacteal feeding [21,22,50]. We found that women who gave birth at home or in the village were three times as likely to practice prelacteal feeding compared to those who gave birth in a health facility. Women who give birth at home or in villages may not receive skilled birth attendant supervision, leading to poor newborn and postnatal guidance and care. Additionally, traditional birth attendants may have influenced women to introduce prelacteal feeds instead of initiating early breastfeeding [22,25]. Increasing awareness about health facility births, particularly among disadvantaged and rural women, is necessary to optimise exclusive breastfeeding.

## Conclusion

Our study revealed that prelacteal feeding is practised among women in PNG, implying that exclusive breastfeeding behaviours are suboptimal. Women who were less educated, had a cesarean section, had home or village births, and lived in the Islands region were more likely to provide prelacteal feeds. Women and, ideally, families more generally, must have access to effective health information and educational resources that will increase their understanding of infant and young child feeding. This study calls for the strengthening of healthcare providers’ capacity to encourage more health facility births and improve mothers’ and household awareness of exclusive breastfeeding while addressing misconceptions and other barriers to breastfeeding to reduce prelacteal feeding. Furthermore, breastfeeding counselling and peer support for exclusive breastfeeding at antenatal clinics and outreach healthcare services should be included as an integral component of breastfeeding promotion programs in PNG and other similar settings.

## Data Availability

All data produced are available online at The DHS Program website: https://dhsprogram.com/data/available-datasets.cfm.

## Acknowledgements

We are thankful to the ICF International and DHS Program for permitting us to use the 2016– 2018 PNGDHS dataset to conduct this study.

## Supplementary materials

STROBE Checklist

## Contributors

MM conceptualized and designed the study. MM and EN conducted statistical analyses. MM drafted the original manuscript. MM, EN and GG contributed to the writing and editing of the manuscript. MM and GG reviewed and edited the manuscript. All the authors reviewed and approved the final version of the manuscript. MM is the guarantor and accepts full responsibility for the work.

## Patient consent for publication

Not required.

## Ethics approval

The 2016–2018 PNGDHS protocol was reviewed and approved by the Institutional Review Board (IRB) of ICF and IRB of host country. Informed consent was obtained from all participants before the interviews were conducted. For this study, a formal request to use the raw data was obtained from the DHS program. Permission was granted by the DHS program to access the data, which was anonymized for analysis.

## Provenance and peer review

Not commissioned; externally peer reviewed.

## Data availability statement

The 2016–2018 PNGDHS data are publicly available in a public, open-access repository. Data can be obtained upon a registration-access request from the DHS Program website at https://dhsprogram.com/data/available-datasets.cfm.

